# Final Days with Advanced Heart Failure: Facility Variation in Home Time and Hospice Enrollment

**DOI:** 10.64898/2026.11.24.26368915

**Authors:** Na Ouyang, Ling Han, Erica A. Abel, Eric C. DeRycke, Daniel Kinder, Laura Moreines, Nancy S. Redeker, Leslie A. Curry, Carol Luhrs, Cari Levy, Jennifer V. Ibarra, Dio Kavalieratos, Kathleen M. Akgun, Larry A. Allen, Shelli L. Feder

## Abstract

**Background:** Days spent at home and hospice enrollment describe different but potentially related aspects of high-quality end-of-life care for people with advanced heart failure (aHF). However, how much variation in these quality outcomes exists and what drives between-facility differences is unknown.

**Methods:** This retrospective cohort study included people with aHF at 119 Veterans Affairs medical centers (VAMCs) who died in 2022–2023. Outcomes were days at home during the last 90 days of life and hospice enrollment. Multilevel linear and logistic regression models with facility random intercepts quantified between-facility variation before and after adjustment for patient characteristics, facility complexity, and facility volume.

**Results:** The cohort included 10,114 decedents (mean [standard deviation, SD] age, 76.5 [9.6] years; 98.2% male). Patients spent a mean (SD) of 59.4 (29.9) days at home during their final 90 days of life, and 4521 (44.7%) enrolled in hospice. For days at home, the adjusted intraclass correlation coefficient (ICC) was 0.95%. The adjusted median difference between two randomly selected VAMCs was 2.7 days (95% CI, 2.0–3.7). Measured patient and facility characteristics explained 29% of the between-facility variance. For hospice enrollment, the adjusted ICC was 3.6%. The adjusted median odds ratio was 1.40 (95% CI, 1.31–1.51), indicating a median 40% higher odds of hospice use when comparing two similar patients treated at two randomly selected VAMCs. Between-facility variance increased 15.0% after adjustment for patient characteristics, facility complexity, and volume.

**Conclusions:** People with aHF spent one-third of their final 90 days of life in institutional settings; fewer than half received hospice. Adjusted home time differed little across VAMCs, while hospice enrollment varied substantially. Persistent variation in hospice enrollment suggests that interventions targeting facility-level care processes, such as referral practices, could improve hospice utilization among people with aHF.

**Clinical Perspective:** *What is new?:*

- In this national VA-based cohort, people with advanced heart failure spent on average 59.4 of their final 90 days of life at home, and 44.7% enrolled in hospice.
- Hospice enrollment, but not time at home, varied widely across 119 medical centers (median odds ratio 1.40). This variation persisted after accounting for patient and medical center characteristics.

*What are the clinical implications?:*

- Fewer than half of people with advanced heart failure enroll in hospice, highlighting missed opportunities for end-of-life support.
- Medical center–level practices, beyond patient or facility characteristics, may shape access to hospice and represent opportunities for quality improvement.

## INTRODUCTION

Heart failure (HF) affects approximately 7.7 million US adults and was listed as an underlying or contributing cause of 14.6% of all US deaths in 2023.^1^ People with advanced HF (aHF; stage D) experience symptoms that interfere with daily life and recurrent hospitalizations despite attempts to optimize guideline-directed medical therapy.^2^ Many people with HF prefer to remain at home as they approach the end of life (EOL), close to family and familiar surroundings;^3^ however, 83% of patients with HF were hospitalized during their final year of life.^4^ Hospice has been associated with better symptom management and lower acute care use,^5^ but its utilization remains limited among this population.^6^

Both days spent at home and hospice enrollment are increasingly recognized as indicators of high-quality EOL care, as they reflect outcomes that many patients and families prioritize: avoiding burdensome hospital-based care and spending their remaining time at home with adequate symptom management and support.^7–9^ Previous studies evaluated home time after HF hospitalizations,^10,11^ but data on time at home during the final months of life among people with aHF remains limited. Hospice utilization varies significantly across hospitals.^6^ This variation may arise from patient case mix, facility resources, staffing, and care processes. Prior studies have reported differences in hospice use by patient age, race and ethnicity, insurance coverage, hospital location, and teaching status.^6,12^ However, the extent to which patient and facility characteristics explain between-facility variation in EOL days at home and hospice enrollment remains unclear. Persistent variation after adjustment for patient case mix and facility resources may identify opportunities to improve care processes, such as hospice referral and care coordination.

To address these gaps, we examined EOL days at home and hospice enrollment among people with aHF in the Veterans Health Administration (VA), the largest integrated healthcare system in the US. We aimed to quantify between-facility variation in these outcomes and assess how much variation is explained by measured patient and facility characteristics. These findings may identify gaps in EOL care and inform the development of interventions to increase time at home and improve hospice access for people with aHF.

## METHODS

### Study Design and Data Sources

We conducted a retrospective observational study using the HEART-PAL cohort, which combines electronic health record data from the VA Corporate Data Warehouse and the Centers for Medicare and Medicaid Services. We followed the REporting of studies Conducted using Observational Routinely-collected health Data (RECORD) guideline, an extension of the Strengthening the Reporting of Observational Studies in Epidemiology (STROBE) guidelines.^13^ The VA Connecticut Healthcare System Institutional Review Board approved the study and waived the requirement for informed consent.

### Study Population

The HEART-PAL cohort has been described previously and used to examine variation across Veterans Affairs Medical Centers (VAMCs) in specialist palliative care and its associations with inpatient hospice use and family-reported EOL care quality.^14–18^ Briefly, people with aHF were identified during 2017 through 2022 using a validated administrative algorithm (Table S1).^19^ For this study, we included the cohort members who died between January 1, 2022, and December 31, 2023. Patients were assigned to the VAMC where they received the largest proportion of their care during the study period. Analyses were restricted to VAMCs with at least 20 eligible decedents to improve the stability of facility estimates.

### Outcomes

The outcomes were home time and hospice enrollment. Home time was defined as the number of days in the 90 days preceding death when a patient was not in an institutional care setting, including acute care hospitals, inpatient rehabilitation facilities, skilled nursing facilities, nursing homes, and inpatient hospice units.^11,20,21^ Each day was counted only once when records from different institutional settings overlapped. Values ranged from 0 to 90 days, with higher values indicating more time outside institutional settings. Because nursing homes may serve as a patient’s usual residence and definitions of home time vary in the classification of nursing-facility days,^22^ we conducted a sensitivity analysis using an alternative definition in which nursing home days were counted as days at home.

We identified hospice enrollment using VA-specific treating specialty codes, stop codes, Fee Event codes, and claims from the VA Cohort hospice files. Patients were considered enrolled in hospice at the first qualifying hospice record, which included hospice enrollment, an inpatient hospice stay, or another qualifying hospice encounter.

### Covariates

We collected covariates from one year before the study observation window. Patient-level covariates included age, sex, marital status, dual Medicare-Medicaid eligibility, VA service-connected disability status, VA enrollment priority group, Elixhauser Comorbidity Index, number of chronic medications, number of emergency department visits, the JEN Frailty Index (JFI), and the VA Care, Assessment, Need (CAN) risk score.^23,24^ We included cohort entry year as a fixed covariate to account for secular trends across the 2017–2022 identification window. We categorized the JFI using established thresholds for low, moderate, and high risk of long-term institutionalization. We categorized CAN scores using VA operational thresholds, including the 60th and 90th percentiles of predicted one-year hospitalization or mortality risk.

Facility characteristics included facility complexity and volume. Facility complexity described the level of services provided in a VAMC, grouped into high complexity (levels 1a, 1b, and 1c), medium complexity (level 2), and low complexity (level 3). This standardized VA administrative classification incorporates patient volume and clinical risk; clinical service complexity, including ICU and surgical capabilities, complex clinical programs (e.g., polytrauma care, rural facility), community care; and teaching and research activity (e.g., residency training and research funding).^25,26^ Facility volume was defined as the number of eligible decedents with aHF attributed to each VAMC and was log-transformed for regression analyses due to its right-skewed distribution.

### Statistical Analysis

We summarized categorical variables using frequencies and percentages and continuous variables using means and standard deviations (SD). We fitted linear and logistic mixed-effects regression models for days at home and hospice enrollment, respectively, with a facility random intercept to account for clustering of patients within facilities. For each outcome, we fitted three sequential models: an intercept-only model to establish baseline variance, a model adjusted for patient characteristics, and a fully adjusted model that additionally included facility complexity and log-transformed facility volume. We assessed multicollinearity among covariates using variance inflation factors prior to model fitting; all values were below conventional thresholds for concern. We calculated intraclass correlation coefficients (ICCs) for each model to quantify the proportion of outcome variation attributable to facilities. We calculated proportional changes in estimated between-facility variance for each adjusted model relative to the null model. For hospice enrollment, we calculated the adjusted median odds ratio (aMOR) to summarize residual between-facility variation as the median difference in odds between two similar patients treated at two randomly selected VAMCs.

From the fully adjusted models, we generated patient-level predictions that incorporated the random effect for each patient’s VAMC and averaged these predictions within facilities to estimate adjusted mean days at home and rates of hospice enrollment. Waterfall plots displayed these facility estimates in rank order and by facility complexity. We classified VAMCs as above, below, or not significantly different from the national mean based on whether their bootstrapped 95% confidence intervals (CIs) fell entirely above, entirely below, or included the national mean, respectively. We also estimated additional and deficit days at home and hospice enrollment relative to national averages to describe the population-level magnitude of facility differences.

For the home-days models, we examined residual distributions to assess model assumptions and used R² and root mean squared error (RMSE) to evaluate model fit. For the hospice model, we assessed discrimination using the C-statistic and calibration by comparing observed and predicted probabilities.

We conducted three sensitivity analyses. First, we repeated the days-at-home analyses using the alternative definition described above. Second, we compared each VAMC with facilities in the same complexity tier. Third, to assess whether a high-volume VAMC disproportionately influenced each tier-specific reference, we recalculated the reference means after excluding the VAMC with the most eligible decedents in each tier. We excluded these VAMCs only from the reference-mean calculation and retained all patients and facilities in outcome comparisons. We compared aggregate days at home and hospice enrollment above and below expected under the original and recalculated peer references.

We used two-sided tests and considered p<.05 statistically significant. We examined missingness for each outcome and model covariate and used complete cases for adjusted analyses. Analyses were conducted using SAS, version 9.4 (SAS Institute).

## RESULTS

### Sample Characteristics

Of 140 VAMCs in the HEART-PAL cohort, we excluded 21 with fewer than 20 eligible decedents, yielding 10,114 decedents with aHF across 119 VAMCs (Table 1). The sample’s mean age was 76.5 years (SD, 9.6); most were male (98.2%), White (66.3%), and non-Hispanic (95.3%). Of the sample, 46.7% were rated as having high disability, and 81.7% had CAN scores above 90, indicating they were in the top 10% of risk for hospitalization or death within the next year. Mean Elixhauser scores were 8.1 (SD, 3.2). Facility volume ranged from 21 to 246 (median, 77; interquartile range, 47–109). Facilities included 84 high-complexity, 20 medium-complexity, and 15 low-complexity VAMCs.

**Table 1:** Patient Characteristics by Days at Home and Hospice Enrollment.

| **Variables** | **Total Sample** |  | **Days at Home in Last 90 Days** |  | **Hospice Enrollment** |  |  |
|---|---|---|---|---|---|---|---|
|  |  |  |  |  | **No Hospice (N=5593)** | **Hospice Enrollment (N=4521)** |  |
|  | **N** | **Mean (SD) or %** | **Mean (SD) or r** | **p-value** | **Mean (SD) or N (%)** | **Mean (SD) or N (%)** | **p-value** |
| Age (Years) | 10114 | 76.5 (9.6) | 0.02<sup>*</sup> | .024 | 74.5 (9.5) | 78.8 (9.0) | <.0001 |
| Gender |  |  |  | .021 |  |  | .812 |
| Male | 9936 | 98.2% | 59.5 (29.9) |  | 5493 (55.3%) | 4443 (44.7%) |  |
| Female | 178 | 1.8% | 54.3 (31.2) |  | 100 (56.2%) | 78 (43.8%) |  |
| Race (proportion, %) |  |  |  | <.0001 |  |  | <.0001 |
| White | 6706 | 66.3% | 60.5 (29.5) |  | 3450 (51.5%) | 3256 (48.6%) |  |
| Black | 2551 | 25.2% | 57.1 (30.5) |  | 1652 (64.8%) | 899 (35.2%) |  |
| American Indian or Alaska Native | 57 | 0.6% | 48.3 (32.3) |  | 28 (49.1%) | 29 (50.9%) |  |
| Asian | 36 | 0.4% | 57.4 (33.6) |  | 25 (69.4%) | 11 (30.6%) |  |
| Native Hawaiian or Other Pacific Islander | 78 | 0.8% | 61.9 (25.7) |  | 49 (62.8%) | 29 (37.2%) |  |
| Mixed | 81 | 0.8% | 57.1 (32.2) |  | 54 (66.7%) | 27 (33.3%) |  |
| Unknown | 605 | 6.0% | 57.9 (30.8) |  | 335 (55.4%) | 270 (44.6%) |  |
| Ethnicity |  |  |  | .037 |  |  | .570 |
| Non-Hispanic | 9642 | 95.3% | 59.3 (30.0) |  | 5326 (55.2%) | 4316 (44.8%) |  |
| Hispanic | 472 | 4.7% | 62.2 (28.3) |  | 267 (56.6%) | 205 (43.4%) |  |
| Marital status (proportion, %) |  |  |  | <.0001 |  |  | <.0001 |
| Married | 5145 | 50.9% | 62.6 (28.0) |  | 2674 (52.0%) | 2471 (48.0%) |  |
| Divorced/Separated/Widowed | 3799 | 37.6% | 55.9 (31.5) |  | 2365 (62.3%) | 1434 (37.8%) |  |
| Single/Unknown/Missing | 1170 | 11.6% | 56.9 (31.1) |  | 554 (47.4%) | 616 (52.7%) |  |
| Medicare (proportion, %) |  |  |  | <.0001 |  |  | <.0001 |
| No Medicare | 334 | 3.3% | 63.1 (28.7) |  | 264 (79.0%) | 70 (21.0%) |  |
| Medicare only | 8222 | 81.3% | 61.2 (28.6) |  | 4382 (53.3%) | 3840 (46.7%) |  |
| Dual Eligible | 1558 | 15.4% | 49.3 (34.6) |  | 947 (60.8%) | 611 (39.2%) |  |
| Disability (proportion, %) |  |  |  | <.0001 |  |  | .0004 |
| High | 4725 | 46.7% | 47.7 (30.6) |  | 2625 (55.6%) | 2100 (44.4%) |  |
| Low-moderate | 1451 | 14.3% | 61.6 (28.6) |  | 779 (53.7%) | 672 (46.3%) |  |
| Low income | 2484 | 24.6% | 59.1 (30.1) |  | 1439 (57.9%) | 1045 (42.1%) |  |
| No service connection | 1256 | 12.4% | 64.7 (27.0) |  | 635 (50.6%) | 621 (49.4%) |  |
| Unknown/Missing | 198 | 2.0% | 55.8 (33.1) |  | 115 (58.1%) | 83 (41.9%) |  |
| Elixhauser Index | 10114 | 8.1 (3.2) | -0.15<sup>*</sup> | <.0001 | 8.0 (3.2) | 8.2 (3.2) | .001 |
| Number of medications | 10114 | 9.2 (4.2) | 0.00<sup>*</sup> | .875 | 9.3 (4.2) | 8.9 (4.3) | <.0001 |
| ED visits in the past year | 10114 | 4.7 (4.0) | -0.08<sup>*</sup> | <.0001 | 4.6 (3.7) | 4.7 (4.4) | .227 |
| JEN Frailty Index | 10114 | 6.6 (2.2) | -0.16<sup>*</sup> | <.0001 | 6.5 (2.2) | 6.6 (2.2) | .015 |
| ICU admissions total past year | 10114 | 0.7 (1.1) | -0.04<sup>*</sup> | <.0001 | 0.7 (1.1) | 0.7 (1.1) | .876 |
| Hospital Admissions in the Past Year | 10114 | 3.8 (2.3) | -0.11<sup>*</sup> | <.0001 | 3.8 (2.5) | 3.8 (2.1) | .951 |
| 1-Year Mortality/Hospitalization Risk Category (CAN score) (proportion, %) |  |  |  | <.0001 |  |  | <.0001 |
| CAN score <=60 | 33 | 0.3% | 63.7 (28.6) |  | Suppressed | Suppressed |  |
| 60< CAN score <=90 | 1635 | 16.2% | 64.8 (27.9) |  | 1082 (66.2%) | 553 (33.8%) |  |
| CAN score >90 | 8268 | 81.7% | 59.1 (29.7) |  | 4396 (53.2%) | 3872 (46.8%) |  |
| Unknown/Missing | 178 | 1.8% | 26.2 (34.6) |  | 91 (51.1%) | 87 (48.9%) |  |
| Cohort Entry Year (proportion, %) |  |  |  | .079 |  |  | .005 |
| 2017 | 899 | 8.9% | 60.9 (29.9) |  | 519 (57.7%) | 380 (42.3%) |  |
| 2018 | 1201 | 11.9% | 58.9 (30.9) |  | 693 (57.7%) | 508 (42.3%) |  |
| 2019 | 1589 | 15.7% | 60.4 (30.4) |  | 917 (57.7%) | 672 (42.3%) |  |
| 2020 | 1607 | 15.9% | 59.5 (30.3) |  | 887 (55.2%) | 720 (44.8%) |  |
| 2021 | 2470 | 24.4% | 59.8 (30.1) |  | 1352 (54.7%) | 1118 (45.3%) |  |
| 2022 | 2343 | 23.2% | 58.0 (28.7) |  | 1222 (52.2%) | 1121 (47.8%) |  |
| 2023 | Suppressed | Suppressed | 45.4 (28.8)) |  | Suppressed | Suppressed |  |
| Facility Complexity |  |  |  | .413 |  |  | .503 |
| 1 | 8743 | 86.5% | 59.3 (29.9) |  | 4851 (55.5%) | 3892 (44.5%) |  |
| 2 | 911 | 9.0% | 60.6 (30.2) |  | 487 (53.5%) | 424 (46.5%) |  |
| 3 | 460 | 4.5% | 59.8 (30.8) |  | 255(55.4%) | 205 (44.6%) |  |
| Urban/Rural/Highly Rural |  |  |  | <.0001 |  |  | .132 |
| Highly Rural or Isolated | 84 | 0.8% | 55.8 (31.7) |  | 43 (51.8%) | 40 (48.2%) |  |
| Rural | 2841 | 28.1% | 61.9 (29.2) |  | 1528 (53.8%) | 1313 (46.2%) |  |
| Urban | 7187 | 71.1% | 58.5 (30.2) |  | 4020 (55.9%) | 3167 (44.1%) |  |
| Days at Home in Last 90 Days | 10114 | 59.4 (29.9) | – | – | – | – | – |
| Hospice Enrollment | 4521 | 44.7% | – | – | – | – | – |

*Note:* <sup>\*</sup> correlation coefficient from Pearson or Spearman correlation analysis.
*Abbreviation:* Elixhauser: Elixhauser Comorbidity Index (calculated from ICD-10 codes from inpatient and outpatient visits up to 12 months before cohort entry); CAN score: Care, Assessment, Needs (CAN) score, a one-year mortality risk score calculated from patient demographic, clinical, vital sign, medication, and utilization features and is updated weekly; ED visits: emergency department visits; JFI VA Score: JEN Frailty Index, representing impairments and multimorbid clusters with high long-term institutionalization risk.
*Suppressed:* Counts below 11 and their corresponding percentages were suppressed in accordance with data use requirements. Additional cells were suppressed to prevent calculation of suppressed counts.

### Crude and Risk-Adjusted Home Time

Patients with aHF spent, on average, 59.4 of the last 90 days of life at home (SD, 29.9). Across VAMCs, the mean observed facility-level home time was 59.5 days (SD, 5.29; range, 42.6–72.1 days). In the fully adjusted model, facility-level risk-standardized means ranged from 55.6 to 63.0 days (Figure 1, panels A and B; Table S2). After adjustment, unmarried status, dual eligibility, greater comorbidity and frailty, and prior emergency department use were associated with fewer days at home (Table 2).

**Figure 1.**
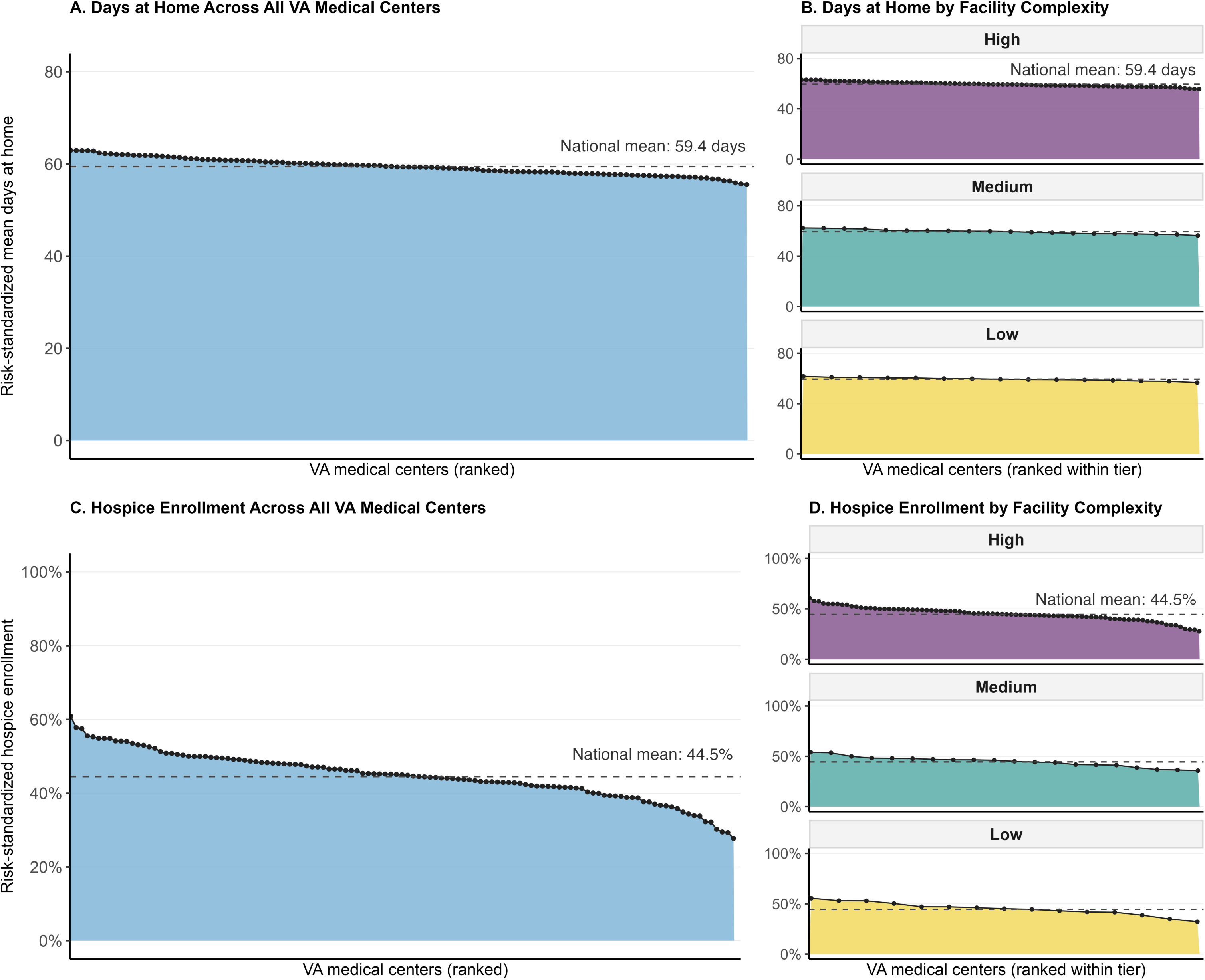
Risk Standardized Days at Home and Hospice Enrollment Across VAMCs. Panel A, Risk-standardized mean days at home across facilities. Panel B, Days at home by facility complexity. Panel C, Risk-standardized rate of hospice enrollment across facilities. Panel D, Hospice enrollment by facility complexity. Centers are displayed from higher to lower outcome values, overall or within complexity tier. Each point represents a VA medical center (VAMC). Dashed lines indicate the national means (59.4 days at home and a hospice enrollment rate of 44.5%). Complexity tiers are high (1), medium (2), and low (3).

**Table 2:** Casemix Variables in Fully Adjusted Model.

| Covariate | Days at Home in the Last 90 Days |  | Hospice Enrollment |  |
|---|---|---|---|---|
|  | β (95% CI) | P-value | OR (95% CI) | P-value |
| **Intercept** | 74.23 (60.43–88.03) | <.0001 | 0.01 (0.00-0.02) | <.0001 |
| **Age** | 0.02 (−0.05–0.10) | .580 | 1.05 (1.04–1.05) | <.0001 |
| **Married Status - Married** | – | – | 1.000 | – |
| Divorced/Separated/Widowed | −5.13 (−6.35 – −3.91) | <.0001 | 0.87 (0.79–0.96) | .005 |
| Single/Unknown/Missing | −4.65 (−6.38 – −2.92) | <.0001 | 1.02 (0.89–1.16) | .823 |
| **Medicare Eligibility - No Medicare** | – | – | 1.000 | – |
| Medicare only | −2.18 (−5.39–1.02) | .182 | 1.54 (1.16–2.05) | .003 |
| Dual Eligible | −12.68 (−16.23 – −9.13) | <.0001 | 1.40 (1.04–1.89) | .028 |
| **Elixhauser** | −0.60 (−0.79–−0.41) | <.0001 | 1.03 (1.02–1.05) | <.0001 |
| **Number of Drug Classes** | 0.44 (0.29–0.59) | <.0001 | 0.98 (0.97–0.99) | .002 |
| **Disability - High** | – | – | 1.000 | – |
| Low-moderate | 4.21 (2.64–5.78) | <.0001 | 1.02 (0.90–1.15) | .794 |
| Low income | 4.38 (2.67–6.10) | <.0001 | 0.96 (0.86–1.07) | .450 |
| No service connection | 5.67 (3.78–7.56) | <.0001 | 1.03 (0.90–1.18) | .685 |
| Unknown/Missing | 1.97 (−2.06–5.99) | .337 | 0.94 (0.69–1.27) | .673 |
| **CAN Score <=60** | – | – | 1.000 | – |
| 60< CAN score <=90 | 1.27 (−8.32–10.83) | .795 | 0.79 (0.35–1.78) | .562 |
| CAN score >90 | −3.10 (−12.57–6.37) | .521 | 0.96 (0.42–2.17) | .921 |
| Unknown/Missing | −31.63 (−42.62 to −20.64) | <.0001 | 0.97 (0.41–2.31) | .941 |
| **ED Visits in Prior Year** | −0.22 (−0.39–−0.05) | .013 | 1.01 (1.00–1.02) | .039 |
| **JFI VA Score** | −1.55 (−1.87–−1.22) | <.0001 | 1.01 (0.99–1.03) | .383 |
| **Cohort Entry Year** | 0.57 (−1.31–2.46) | .551 | 0.94 (0.81–1.08) | .374 |
| **ICU Admissions in Prior Year** | −0.12 (−0.62–0.37) | .619 | 0.99 (0.95–1.03) | .651 |
| **Facility Complexity = 3** | – | – | 1.000 | – |
| Facility Complexity =1 | −1.43 (−5.45–2.60) | .483 | 0.96 (0.65, 1.39) | .811 |
| Facility Complexity =2 | 0.09 (−3.90–4.08) | .964 | 1.18 (0.83, 1.67) | .348 |
| **Log Number of Decedents** | 0.77 (−1.23–2.78) | .447 | 1.19 (0.96, 1.48) | .105 |

*Abbreviation:* Elixhauser: Elixhauser Comorbidity Index (calculated from ICD-10 codes from inpatient and outpatient visits up to 12 months before cohort entry); CAN score: Care, Assessment, Needs (CAN) score, a one-year mortality risk score calculated from patient demographic, clinical, vital sign, medication, and utilization features and is updated weekly; ED visits: emergency department visits; JFI VA Score: JEN Frailty Index, representing impairments and multimorbid clusters with high long-term institutionalization risk.

### Crude and Risk-Adjusted Rate of Hospice Enrollment

Overall, 44.7% of patients with aHF were enrolled in hospice. Across VAMCs, the mean observed facility-level rate of hospice enrollment was 44.5% (SD, 10.4%; range, 13.5%–69.1%). In the fully adjusted model, facility-level risk-standardized rates varied from 27.7% to 60.9% (Figure 1, panels C and D; Table S3). After adjustment, hospice enrollment was more common among older patients and those with Medicare eligibility, greater comorbidity, or prior emergency department use and less common among divorced/separated/widowed patients (Table 2).

### Between-Facility Variation

In the null model, between-facility variance in home time was 11.0 days^2^ (ICC, 1.23%). Adjustment for patient characteristics reduced this variance by 32.0%, to 7.48 days^2^. Adding facility complexity and volume did not further reduce variance. The variance in the fully adjusted model was 7.81 days^2^ (ICC, 0.95%), representing a 29.0% reduction relative to the null model. The median difference in home time between two similar patients at two randomly selected VAMCs was 2.7 days (95% CI, 2.0–3.7). The fully adjusted model explained 9.9% of the variance in observed home time (R², 0.10; RMSE, 28.4 days).

For hospice enrollment, the null-model between-facility variance was 0.107 (ICC, 3.2%). In the fully adjusted model, between-facility variance was 0.123 (ICC, 3.6%), increasing by 15.0% from the null model after adjustment. The aMOR was 1.40 (95% CI, 1.31–1.51), indicating a median 40% higher odds of hospice use when comparing two similar patients treated at two randomly selected VAMCs. The fully adjusted hospice model had modest discrimination (C-statistic, 0.64; 95% CI, 0.63–0.65) and good calibration, with observed and predicted rates differing by less than 3.5 percentage points across deciles of predicted probability.

### Risk-Standardized Outcome Performance

For days at home, 71 (59.7%) performed not significantly different from the national mean, 27 VAMCs (22.7%) performed better, and 21 (17.6%) performed worse (Figure 2, panel A; Table S2). For hospice enrollment, 40 (33.6%) performed not significantly different from the national mean, 44 (37.0%) performed better, and 35 (29.4%) performed worse (Figure 2, panel B; Table S3).

**Figure 2.**
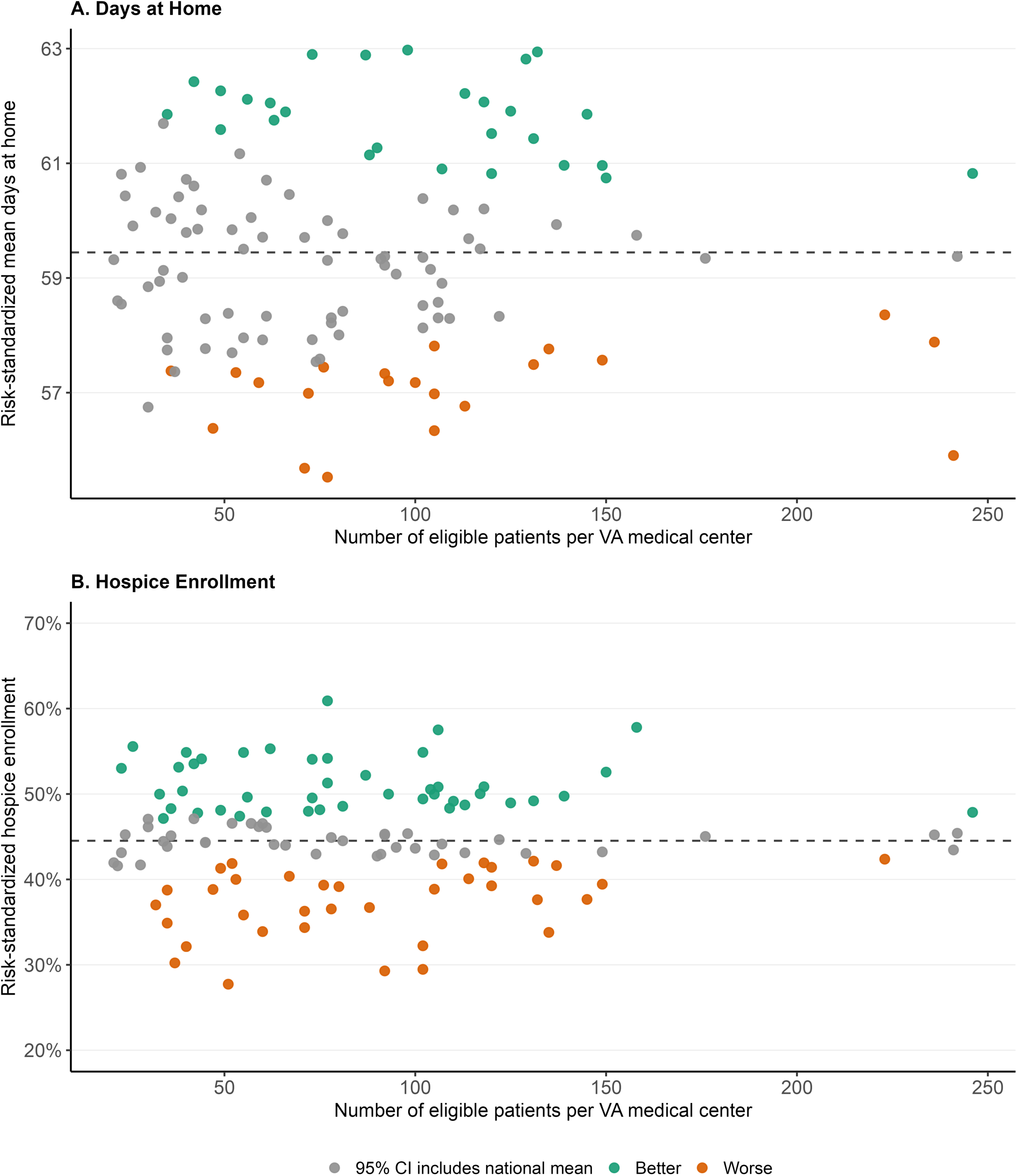
VAMC Classification by Risk-Standardized Days at Home and Hospice Enrollment. Panel A, Days at home. Panel B, Hospice enrollment. Each point represents a VA medical center (VAMC). The x-axis shows the number of eligible patients at that VAMC. Colors reflect whether the bootstrap 95% confidence interval for the risk-standardized outcome lies entirely above the national mean (better), entirely below the national mean (worse), or includes the national mean (as expected). Higher values are classified as better for both outcomes. Dashed lines indicate national means.

### Excess and Deficit Relative to Expected Performance

Facility-level excesses and deficits relative to the national mean totaled 33,720 patient-days at home: 16,817 additional days (2.8% of observed home days) among VAMCs above the national mean and 16,904 fewer days (2.8%) among those below the national mean (Figure 3, panel A). The 10 VAMCs with the largest excesses and the 10 VAMCs with the largest deficits accounted for 45.3% of additional days and 44.3% of deficit days, respectively.

**Figure 3.**
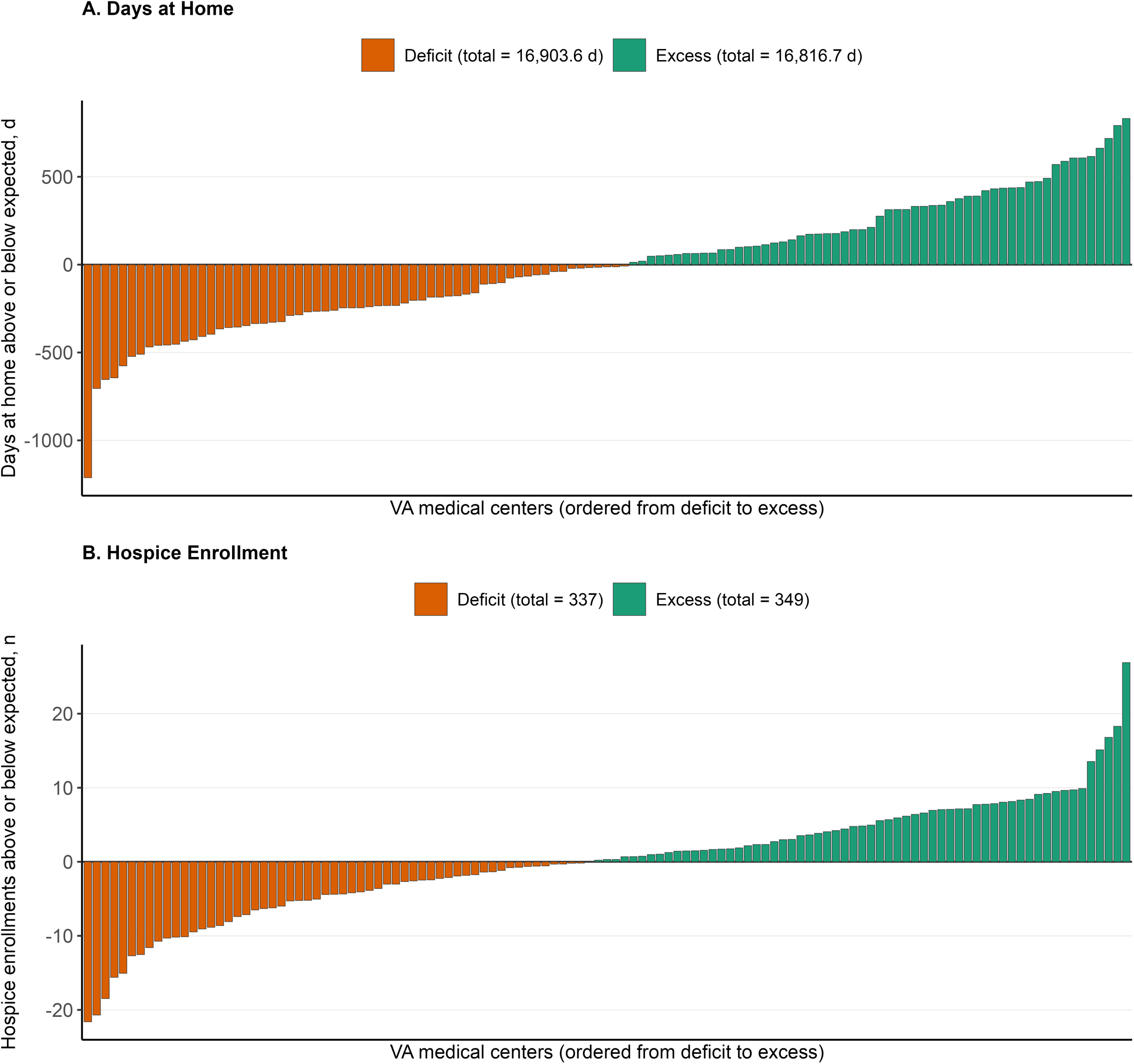
Facility-Level Excesses and Deficits in Days at Home and Hospice Enrollment. Panel A, Days at home above or below expected. Panel B, Hospice enrollments above or below expected. Each bar represents one VA medical center, ordered from the largest deficit to the largest excess. Facility identifiers have been omitted. Negative values indicate deficits and positive values indicate excesses; legend totals are sums across facilities.

Facility-level excesses and deficits relative to the national rate totaled 686 hospice enrollments: 349 additional enrollments (7.7% of observed enrollments) among VAMCs above the national rate and 337 fewer enrollments (7.5%) among those below the national rate (Figure 3, panel B). The 10 VAMCs with the largest excesses and the 10 VAMCs with the largest deficits accounted for 46.4% of additional enrollments and 43.1% of enrollment deficits, respectively.

### Sensitivity Analyses

When nursing home days were counted as home time, patients spent, on average, 69.4 of the last 90 days of life at home (SD, 21.7; facility range, 64.3–73.1). Thirty-one facilities (26.1%) performed not significantly different than the national mean, 36 (30.3%) performed better, and 52 (43.7%) performed worse (Table S4; Figure S1).

Results were similar when each VAMC was evaluated against facilities in the same complexity tier rather than the national mean. Under the primary definition of days at home, days above and below expected represented 1.4% and 1.2% of observed days respectively; both proportions were 1.1% of observed days when nursing home days were counted as home time. Hospice enrollment above and below expected represented 5.7% and 5.3% of observed enrollments, respectively. In leave-one-out analyses, excluding the highest-volume VAMC from each tier changed these estimates by no more than 0.3 percentage points for all outcomes, supporting the stability of the tier-specific reference.

## DISCUSSION

In this national cohort, people with aHF spent one-third of the last 90 days of life in institutional care, and fewer than half enrolled in hospice. Adjusted differences in mean days at home across VAMCs were small, with a median difference of 2.7 days between two randomly selected VAMCs. Measured patient and facility characteristics explained 29.0% of between-facility variance in home time. In contrast, hospice enrollment varied by more than 2-fold across VAMCs even after adjustment for patient characteristics and facility complexity and volume. Low hospice enrollment and substantial residual between-facility variation in hospice use suggest that differences in hospice referral practices and care coordination across VAMCs may be modifiable targets for improving EOL care in aHF.

To our knowledge, this is the first study to provide a national estimate of days at home in the final three months of life among people with aHF. Prolonged institutional care may reflect recurrent hospitalizations, functional decline, and increasing post-acute care needs among people with aHF during EOL.^2,7,27,28^ Prior work has described a cycle in which seriously ill older adults move between hospitals and nursing care facilities during their final months of life, sometimes without returning home or without having prognosis and goals of care adequately addressed.^28^ Although institutional care may be clinically appropriate, substantial time in these settings near the EOL may indicate a mismatch between patients’ care needs, available support, and preferred place of care.^3,28,29^

Adjusted mean days at home varied little across VAMCs, ranging from 55.6 to 63.0 days, a difference of less than 10 days. Most variation occurred among patients within facilities. In contrast, a previous study found home time after HF hospitalizations varied across hospitals, supporting its potential use as a hospital performance measure.^11^ These findings suggest that the usefulness of home time for comparing facility performance after HF hospitalization may not extend to the final months of life. Prior HF research has linked shorter home time to patients’ comorbid conditions.^10^ Other studies have identified caregiver support and access to home-based services as important for meeting patients’ care needs at home.^9,30^ Together, these findings suggest that efforts to improve home-based EOL care should match services to patients’ clinical needs and preferences, and ensure that caregivers have the support needed to provide care at home.

Fewer than half of people with aHF in our cohort enrolled in hospice before death, a rate comparable to that reported among Medicare beneficiaries discharged to home health care after HF hospitalization, 48.2% of whom were receiving hospice at death among those who died within 6 months.^27^ Low hospice use in aHF warrants attention because hospice enrollment is associated with lower acute care use, a lower likelihood of in-hospital death, and increased family satisfaction.^5,31^ Previously identified barriers to hospice enrollment in HF include prognostic uncertainty, limited understanding of hospice services, and patients’ reluctance to transition from familiar cardiology care to an unfamiliar hospice team.^32,33^ Many of these barriers stem from how care is delivered rather than from fixed patient characteristics, suggesting they may be amendable to intervention.

Between-facility variation in hospice enrollment persisted after adjustment for patient characteristics and facility resources. Unmeasured care processes may account for this residual variation, including palliative care consultation, care coordination, and cardiology-hospice collaboration.^32,34,35^ The concentration of excess hospice enrollments among a small number of medical centers in our study suggests that higher hospice use is achievable within the same health system. Comparing care delivery processes between VAMCs with the largest excesses and deficits in hospice enrollment relative to expected rates may identify practices, such as standardized approaches to assessing hospice eligibility, conducting goals-of-care discussions, and coordinating referrals, that could be adopted more broadly.

This study has several limitations. First, hospice enrollment was measured as a binary outcome without information on setting, timing, and duration. The substantial differences in hospice enrollment across VAMCs warrant examining whether these aspects also vary by facility. Second, unmeasured caregiver support and patient preferences may contribute to residual between-facility variation. Future studies should examine these factors to assess whether home time and hospice enrollment align with patients’ goals and caregivers’ needs. Third, this cohort consisted predominantly of older male adults, limiting generalizability to other populations. Finally, the observational design and absence of direct measures of hospice referral and care coordination preclude attributing facility variation to specific care processes. Comparing these practices at facilities with higher and lower adjusted enrollment rates could help identify processes associated with the observed differences.

## CONCLUSION

People with aHF spent substantial time away from home near death, and fewer than half received hospice care. Days at home varied little across VAMCs. However, hospice enrollment varied over 2-fold, after adjusting for measured patient and facility characteristics. Low hospice utilization and persistent variation may reflect modifiable organizational factors for quality improvement.

## Author Contributions

Drs Ouyang and Feder had full access to all of the data in the study and take responsibility for the integrity of the data and the accuracy of the data analysis.

*Concept and design:* Ouyang, Han, Feder

*Acquisition, analysis, or interpretation of data:* All authors

*Drafting of the manuscript:* Ouyang, Feder

*Critical review of the manuscript for important intellectual content:* All authors

*Statistical analysis:* Feder, Han

*Administrative, technical, or material support:* Ibarra

*Supervision:* Feder, Redeker, Allen

## Data Availability

Due to US Department of Veterans Affairs (VA) regulations and our ethics agreements, the analytic data sets used for this study are not permitted to leave the VA firewall without a Data Use Agreement. However, VA data are made freely available to researchers with an approved VA study protocol. For more information, please visit https://www.virec.research.va.gov.

## Acknowledgements

The authors acknowledge the use of Claude Sonnet 4.6 for code generation and syntax refinement, and the use of Claude Sonnet 5 to refine the manuscript’s phrasing, improve expression, and correct grammatical errors. After utilizing this tool, the authors reviewed and edited the content as necessary and took full responsibility for the final text and the accuracy of the work.

## Non-standard Abbreviations and Acronyms

aHF: advanced heart failure
HF: heart failure
EOL: end of life
VA: Veterans Health Administration
VAMC: Veterans Affairs Medical Centers
JFI: JEN Frailty Index
CAN: VA Care, Assessment, Need risk score
SD: standard deviation
ICC: intraclass correlation coefficients
aMOR: adjusted median odds ratio
CI: confidence intervals
RMSE: root mean squared error

